# SARS pandemic exposure impaired early childhood development: A lesson for COVID-19

**DOI:** 10.1101/2020.05.12.20099945

**Authors:** Yunfei Fan, Huiyu Wang, Qiong Wu, Xiang Zhou, Yubo Zhou, Bin Wang, Yiqun Han, Tao Xue, Tong Zhu

## Abstract

Social and mental stressors associated with the COVID-19 pandemic may promote long-term effects on child development. However, reports aimed at identifying the relationship between pandemics and child health are limited. We conducted a retrospective study to evaluate the severe acute respiratory syndrome (SARS) pandemic in 2003 and its relationship to child development indicators using a representative sample across China. Our study involved longitudinal measurements of 14,647 children, 36% of whom (n = 5216) were born before or during the SARS pandemic. Cox models were utilized to examine the effects of SARS on preterm birth and four milestones of development: age to (1) walk independently, (2) say a complete sentence, (3) count from 0 to 10, and (4) undress him/herself for urination. Mixed effect models were utilized to associate SARS with birthweight, body weight and height. Our results show that experiencing SARS during early childhood was significantly associated with delayed milestones, with adjusted hazard ratios of 3.17 [95% confidence intervals (CI): 2.71, 3.70], 3.98 (3.50, 4.53), 4.96 (4.48, 5.49), or 5.57 (5.00, 6.20) for walking independently, saying a complete sentence, counting from 0 to 10, and undressing him/herself for urination, respectively. Experiencing SARS was also associated with reduced body weight. This effect was strongest for preschool children [a weight reduction of 4.86 (0.36, 9.35) kg, 5.48 (−0.56, 11.53) kg or 5.09 (−2.12, 12.30) kg for 2, 3, 4 year-olds, respectively]. We did not identify a significant effect of maternal SARS exposure on birthweight or gestational length. Collectively, our results showed that the SARS pandemic was associated with delayed child development and provided epidemiological evidence to support the association between infectious disease epidemics and impaired child health. These results provide a useful framework to investigate and mitigate relevant impacts from the COVID-19 pandemic.

## Introduction

The 2019 coronavirus infectious disease (COVID-19) outbreak caused a global pandemic^1^. Social distancing policies, which have been shown to mitigate the outbreak^2, 3, 4^, were implemented worldwide^5^. The reduced companionship associated with quarantine, isolation, and school closures^6^ can produce social stress^7^, which may adversely affect child health and development^8^. Furthermore, social distancing policies reshape the living environment for children and often reduce opportunities for physical activity and outdoor exposure^6^, which are critical for psychological and physiological development. Feelings of anxiety, depression and stress spread through social media networks^9, 10^ and negatively impact both children and their caregivers.

The impacts of COVID-19-associated social stress on child health are unclear and may persist long after the pandemic. Thus, evaluations of epidemiological relationships between pandemics and child health are needed to inform planning the relevant interventions. Contemporary population studies are necessary to monitor health changes prospectively during and after the COVID-19 pandemic. Furthermore, lessons from previous pandemics can inform specific hypotheses for future studies and guide preliminary preventions.

The Severe Acute Respiratory Syndrome (SARS) outbreak in 2003 was also caused by a coronavirus ^11^. To mitigate SARS, social distancing policies were applied in China, particularly in Beijing and Guangdong^11^. Those policies have greatly influenced the current COVID-19 pandemic response^4^. An understanding of SARS-related impacts on child health will inform studies aimed at evaluating similar impacts of the COVID-19 pandemic. The present study analyzed the China Family Panel Studies (CFPS)^12^ to examine the epidemiological associations between the SARS pandemic and infant and child health indicators, including developmental milestones, body weight, height, birthweight and preterm birth.

## Methods

### Population data

The CFPS constitutes an ongoing effort to monitor the health and economic status of adults and children in China^12^. Using a well-designed multi-stage sampling approach^13^, children (0 – 15 years old) who were born before and after the SARS pandemic (November 2002 – May 2003) were surveyed in 2010, 2011, 2012 and every two years thereafter. Baseline samples were collected from the 25 most populous provinces and included both SARS hotspots (Beijing municipality and Guangdong Province) and non-hotspots (*e.g*., Heilongjiang Province). Experienced interviewers collected demographic information, birth records, physiological development and cognitive development data using a computer-assisted questionnaire. The study has been approved by the institutional review board at Peking University (Approval IRB00001052-14010). In this study, we focused on two indicators of child development: (1) longitudinally-reported body weight and height; and (2) time (in months) to reach a developmental milestone^14^, including (a) walking independently, (b) saying a complete sentence, (c) counting from 1 to 10, and (d) undressing him/herself for urination. We also considered newborn health indicators of gestational length and birthweight both as potential cofounders for the child development and outcome variables. We obtained all records of 14,754 children from CFPS 2010-2016. We excluded six subjects with ambiguous age or those ≥ 16 years old, and 101 subjects with invalid data for any of the eight study outcomes. A total of 14,647 children with 41,732 longitudinal records were selected for study inclusion. We prepared the analyzed datasets for the eight outcome variables separately (to account for variable patterns of missing data), and each subject was included in at least one health outcome model (Table 1).

**Table 1.**
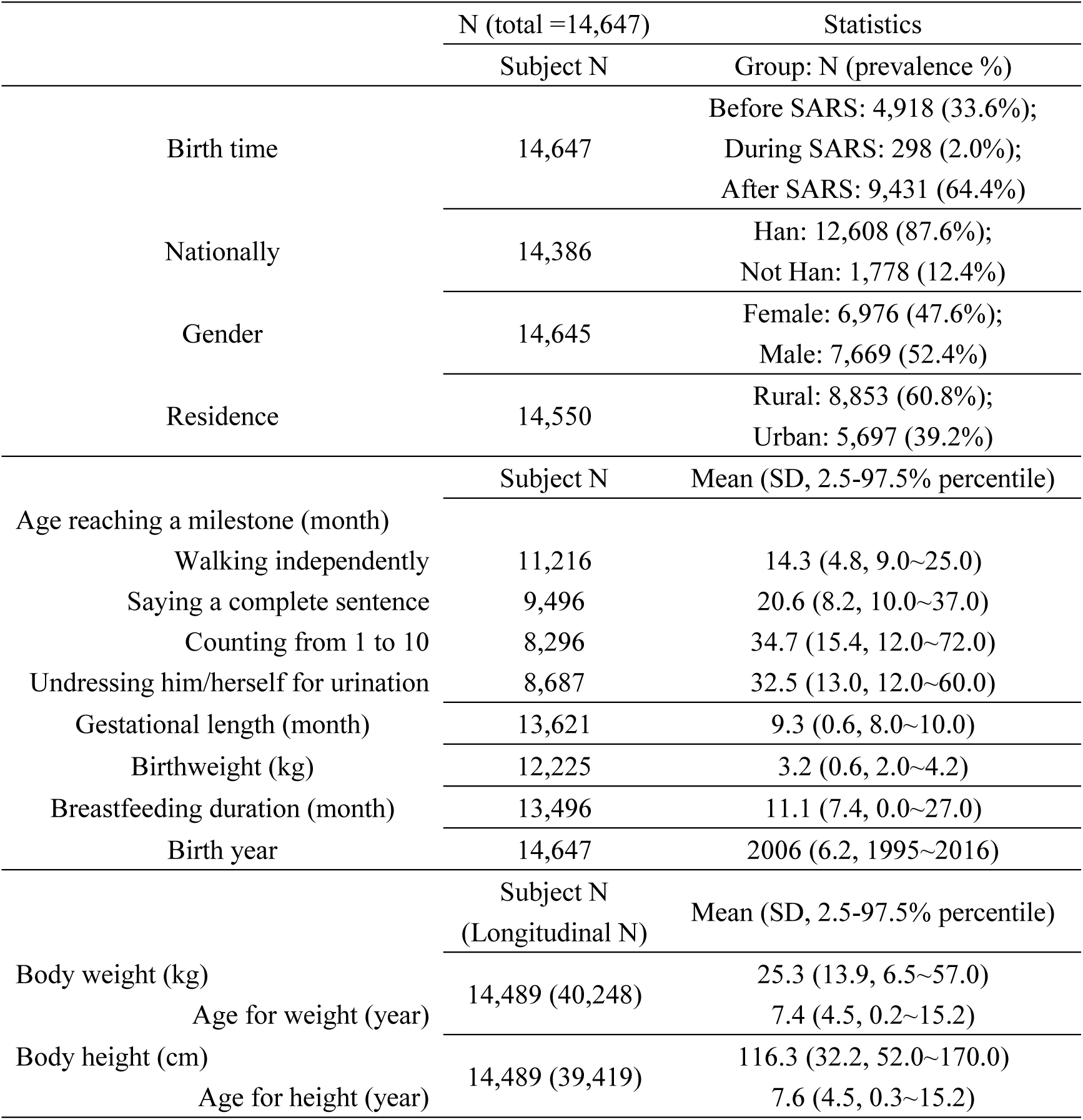
Summary of population characteristics.

### SARS outbreak

The onset of the SARS pandemic occurred November 16^th^, 2002, in Guangdong Province and ended on May 28^th^, 2003^15^. There were 5327 probable cases and 343 deaths. SARS cases were highly clustered in Beijing and Guangdong. In Beijing, there were 2522 cases (47% of total) and the epidemic curve peaked around April 24^th^, 2003; in Guangdong, there were 1504 cases (28%) and the peak occurred around February 8^th^, 2003^15^. To limit SARS transmission, a series of social-distancing policies were implemented. In Beijing, strict quarantine, closing of facilities (*e.g*., shops and schools) and body temperature surveillance were implemented^11^. In the present study, we defined the pandemic period as November 2002 – May 2003, and the disease hotspots as Beijing and Guangdong^15^ (according to the birth province of each child). We assumed that all residents were influenced by the social-distancing policies and that those from hotspot areas experienced greater impacts. Province-level statistics of SARS were obtained from the 2004 China Public Health Statistical Yearbook (http://www.nhc.gov.cn/wjw/tjnj/list.shtml).

### Study design and statistical analyses

We examined the relationship between developmental milestones and exposure to the SARS pandemic. Positive cases included those where birth to milestone-reaching time overlapped with the SARS period. Thus, the pandemic risk during early childhood was characterized by a binary variable, SARS_child_. Similarly, we defined fetal exposure with an additional binary variable, SARS_maternal_. A Cox model regression analysis was conducted to compare time to reach a milestone with exposure indicators (SARS_child_ or SARS_maternal_) after adjusting for several covariates, including residence (urban or rural), sex (female or male), ethnicity, gestation length, birthweight, and breastfeeding status. To control for the nonlinear trend in child health (*e.g*., improvement in nutrition and medical service), we incorporated a spline term with a degree of freedom (DoF) per year; to control for the spatial patterns, we added a random effect term of provincial index to the regression model. Using a similar Cox model, we also associated risk of preterm birth (measured by gestation length) to SARS_maternal_. This model incorporated all of the above covariates, except for gestation length, birthweight, and breastfeeding status.

We used a varying-coefficient model to associate body height or weight with the SARS pandemic, and a binary variable (SARS_child,_*_k_*) to characterize whether a subject was exposed to the SARS pandemic at age *k*. Since all children in the CFPS were affected by SARS at no later than 8 years of age (the oldest child involved in the CFPS was born in 1995), the exposure was coded as a 9-dimensional vector ([SARS_child,_*_0_*, …, SARS_child,_*_8_*]’). We assumed that the effect of SARS varied with developmental stage and applied a varying-coefficient model. In the model, the regression coefficient of SARS was parameterized as a 3-DoF spline function of age (*k*) at the pandemic. Besides the similar set of covariates (residence, sex, nationality, gestation length, birthweight, breastfeeding, temporal nonlinear trend and spatial random term), we also adjusted for a 5-DoF spline term of age at survey and a random term of subject ID. The former term controlled for the change in weight or height with age, and the latter controlled for within-subject correlation of the longitudinal data. A similar regression model with a fixed coefficient was used to associate SARS_maternal_ with birthweight. In the birthweight model, we adjusted for the covariates of residence, sex, nationality, gestation length, temporal nonlinear trend and the spatial random term. For sensitivity analyses, we developed models with mutual adjustments of SARS_child_ and SARS_maternal_ for developmental milestones, body height and weight. Using interaction analyses, we tested whether the estimated effects were the same between hotspot and non-hotspot areas of SARS or among subpopulation strata. Since the associations between pandemic and delayed milestones were estimated according to the longitudinal contrast between generations, we further incorporated spatial contrast within the same generation by examining variations of the effect by pandemic size. We utilized the provincial level number of total cases or deaths as the indicator and examined the interaction between this variable and SARS_child_. All analyses were performed using R statistical software (R-3.4.1). The Cox models were estimated by Package *Survival*, and the varying-coefficient models by Package *DLNM*. To maximize the sample sizes, we used Package *mice* to randomly input missing values in the adjusted covariates for all models.

## Results

### Descriptive statistics

This study involved 14,647 children. Among them, 4,918 (33.6%), 298 (2.0%) and 9,431 (64.4%) were born before, during or after the SARS pandemic, respectively. For all the children born before May 31^st^ of 2003 (n = 4,918 + 298), the mean age at SARS was 4.3 years, with a range of 0 - 8.4 years. Among the 13,621 subjects with valid records of gestation length, 707 (5.2%) children were exposed to SARS prenatally. Given the overlap between the lifespans of CFPS subjects and the SARS pandemic, about one out of three surveyed children were potentially exposed to social stressors during their early childhood or prenatal period. For all available measurements, the mean age to reach the milestones of (a) walking independently, (2) saying a complete sentence, (3) undressing him/herself for urination or (4) counting from 1 to 10 was 14.3 months, 20.6 months, 32.5 months and 36.7 months, respectively. Detailed statistics are displayed in Table 1.

### Developmental milestones

The Cox regression analyses suggested that exposure to SARS during the prenatal stage (SARS_maternal_) or early childhood (SARS_child_) was associated with delayed milestones. Possibly due to the small size of exposed cases, the estimated hazard ratios (HRs) for SARS_maternal_ were weak and their significance levels were sensitive to model settings such as being mutually-adjusted by SARS_child_ or not (Table 2). For SARS_child_, we found strong and robust associations, which were not considerably changed by adjustments for different covariates (Table S1). According to the fully-adjusted models (Table 2), exposure to SARS during early childhood was associated with delayed time to walking independently, saying a complete sentence, counting from 1 to 10, and undressing for urination, with the HR of 3.17 (95% confidence intervals [CI]: 2.71, 3.70), 3.98 (3.50, 4.53), 4.96 (4.48, 5.49), or 5.57 (5.00, 6.20), respectively. Since our exposure indicators were indirect measures of SARS-associated social stress, which could have lasted longer than the pandemic, underestimation of associations is possible due to exposure misclassifications. Simultaneous incorporation of SARS_maternal_ and SARS_child_ into the Cox models improved the accuracy of exposure characterization and may also explain why mutual adjustments enhanced the estimated associations (Table 2). Additionally, we found that the effects of SARS_child_ in pandemic hotspots (Beijing and Guangzhou) were stronger than in non-hotspots, but the between-area difference was statistically significant only for walking (P = 0.032, 0.423, 0.295 or 0.133 for walking, saying, counting or undressing, respectively, Table S2). Using the continuous indicator for pandemic size, we found consistent results (Figure 2 and Table S3). The HR was positively associated with areas with high numbers of SARS cases or deaths (Figure 2). For instance, a 10% increase in the number of SARS deaths was associated with an HR increase of 0.46% (−0.02%, 0.94%), 0.36% (−0.06%, 0.79%), 0.31% (−0.05%, 0.68%) or 0.45% (0.08%, 0.82%) for walking, saying, counting and undressing, respectively (Table S3). We also found that residence in rural areas may enhance the association between SARS_child_ and milestone delay (Table S2).

**Table 2.**
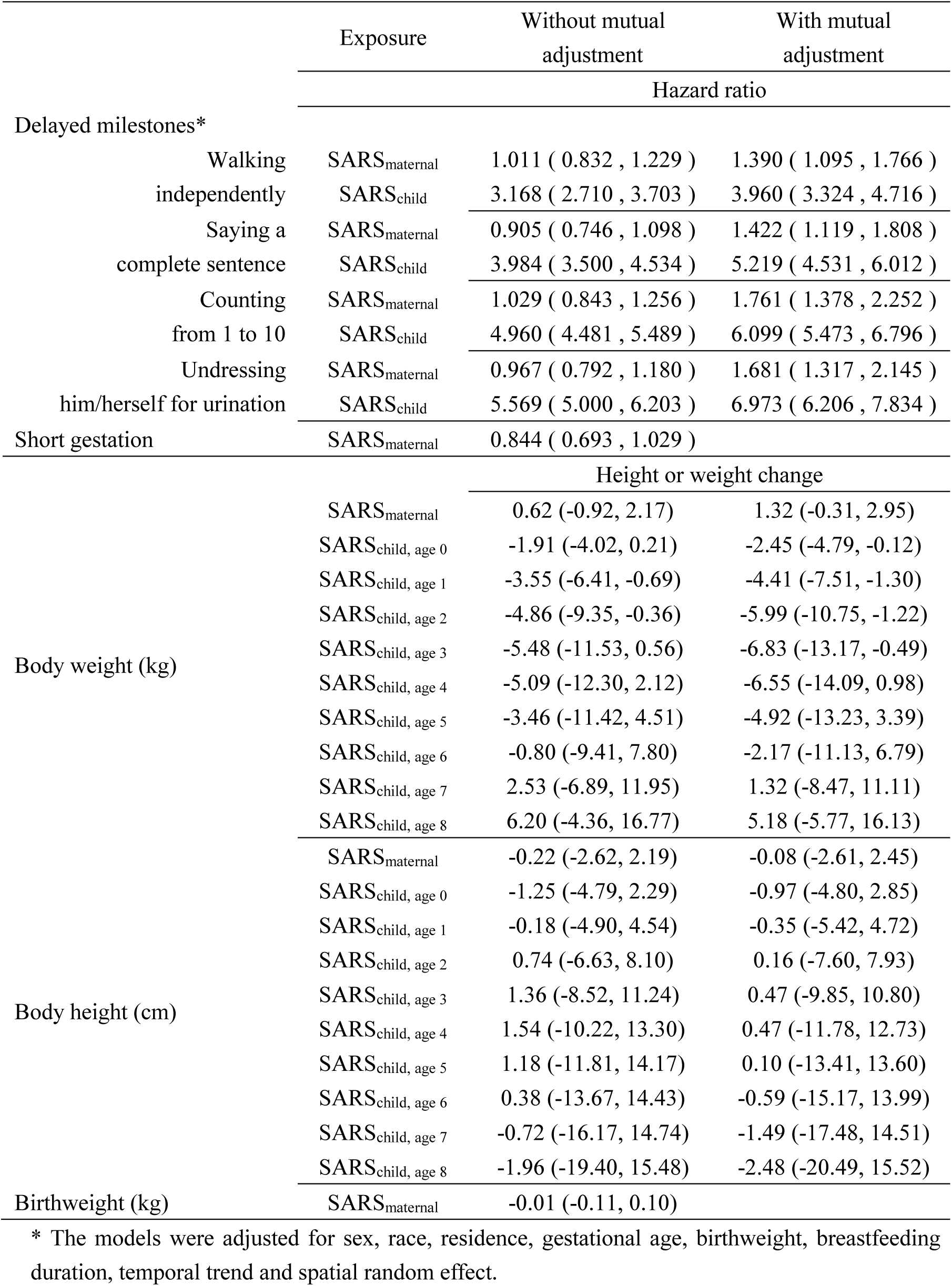
Associations between SARS and indicators of child health.

**Figure 1.**
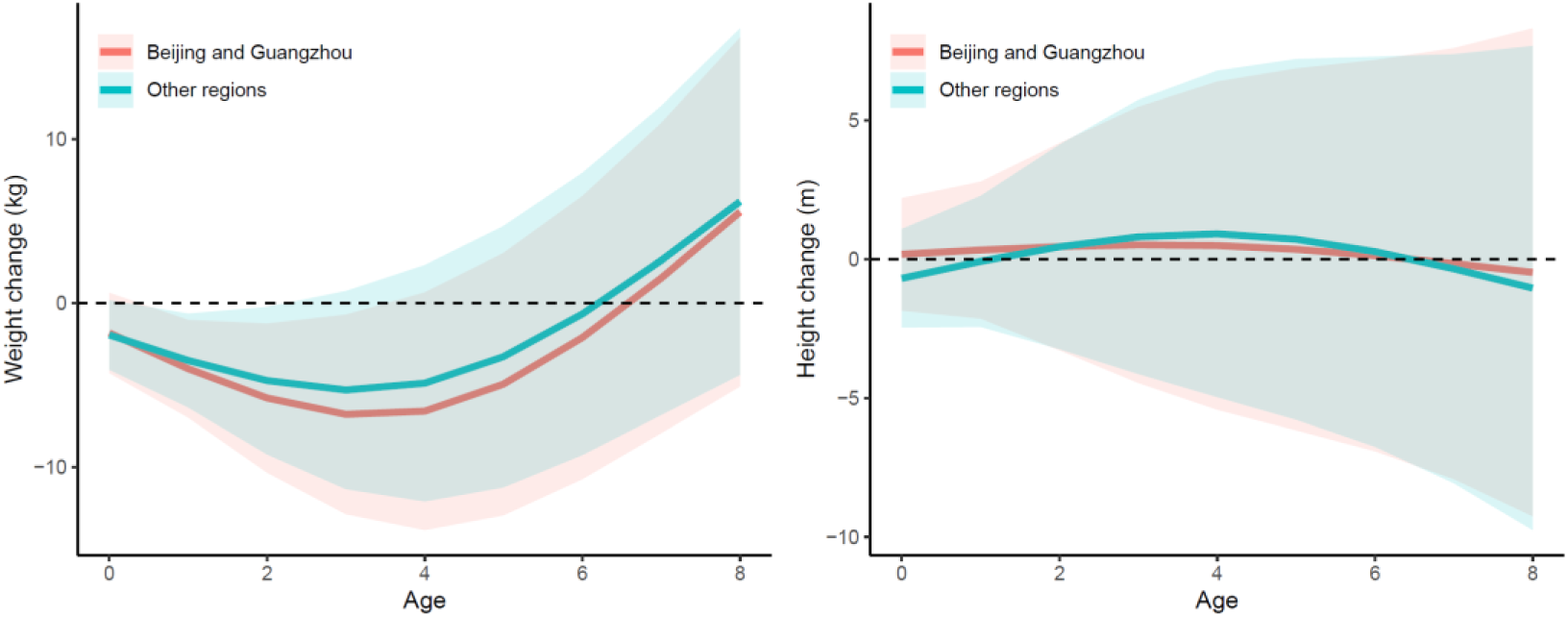
Associations between SARS and body weight (left) and height (right) by age and region. The lines present the estimated associations and the ribbons present the corresponding 95% confidence intervals.

**Figure 2.**
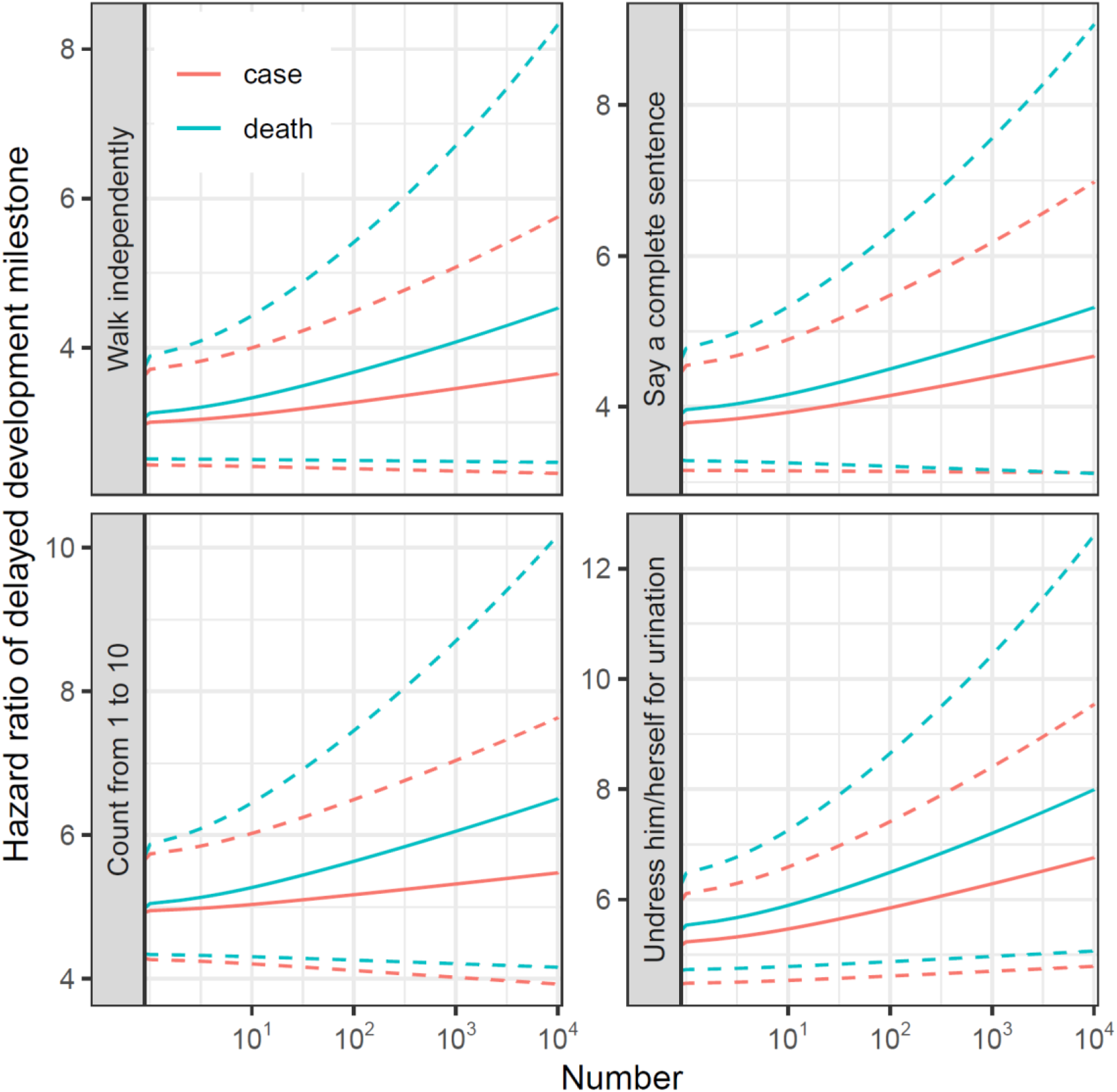
The association between SARS and delayed developmental milestone is enhanced by pandemic size. The pandemic size is indicated by the number of SARS cases (red) or deaths (blue) in a provincial area. The sold lines present how the hazard ratio for an association between SARS and a delayed milestone vary with the pandemic size; and the dashed lines present the 95% confidence intervals.

### Body weight and heigh***t***

We found that exposure to SARS during the early childhood was associated with reduced body weight, but not height. The estimated associations varied with the age during SARS pandemic exposure and peaked at three years of age (Table 2 and Figure 1). According to the fully adjusted model, experiencing SARS at 2-, 3- or 4-years-old was associated with a weight reduction of 4.86 kg (0.36, 9.35), 5.48 kg (0.56, 11.53) or 5.09 kg (−2.12, 12.30), respectively. Similarly, in Beijing and Guangzhou, the effect at 3 years of age was estimated as a body weight reduction of 6.78 kg (0.70, 12.87), which was greater than that observed in other regions (5.30 kg [-0.74, 11.34]; P value = 0.0006 for the likelihood ratio test of the null hypothesis that the overall effect was identical between regions). The association between SARS and body weight reduction might also reflect the pandemic’s effect on delayed development. No significant effects for SARS_maternal_ on weight or height were identified.

### Preterm birth and birthweight

No significant associations between SARS_maternal_ and change in gestational length or birthweight (Table 2) were identified. However, our findings should not be interpreted as evidence against the risk of maternal exposure to SARS. The uncertainty in the models of preterm birth and birthweight may relate to the sample size during the SARS pandemic. For instance, within the exposed period, there were 28 preterm births (gestational length < 37 weeks) and 35 newborns with low birthweight (< 2.5 kg). In addition, our retrospective analysis does not account for fetuses who were severely affected by SARS (*e.g*., prematurely terminated pregnancies). Thus, future prospective studies should evaluate impacts from maternal exposure to the pandemic.

## Discussion

This study presents a retrospective analysis to examine the effect of the SARS pandemic on child health. We found that living through the pandemic during early childhood delayed both physiological and cognitive development. Our results suggest that the current COVID-19 pandemic may have similar impacts on child health. Relevant studies and interventions are urgently needed.

Many biological or behavior pathways indicate that novel infectious disease pandemics, like SARS or COVID-19, can impact child health beyond the effects of infection, through (1) behavioral, (2) environmental, and (3) socioeconomic pathways. First, behavioral changes (wearing masks, quarantines, and reduced outdoor activity) can affect the physiological and psychosocial functions of children. SARS prevention guidelines indicated that masks impair non-verbal communication between children and adults, promoting psychosocial impacts through weakening of social and cognitive connections^16^. Prolonged use of masks can also result in discomfort^17^ and physiological changes^18^, which, for vulnerable individuals, may be particularly harmful. Isolation of children due to infection or exposure may separate children and their caregivers or peers and alter an important learning context for development. For instance, parental absence may impact cognitive achievements^19^ and increase the risk of depression in later life^20^. School closures further reduce companionship, which is an efficient coping measure for social crises^26^. The home-confinement orders increase the prevalence of physical inactivity and sedentary behaviors (*e.g*., watching TV), which are risk factors for cardiovascular diseases, obesity, diabetes, and mental disorders in young people^21^. Second, the pandemic reshapes the living environment of children by keeping them indoors and reducing exposure to the natural environment. According to the biophilia hypothesis^22^, affiliation with nature promotes child developmental psychology^23^. For instance, residential green space has been associated with beneficial effects on cognitive development and brain function in school-aged children^24,25^. Finally, the pandemic is associated with both short- and long-term socioeconomic impacts. During the outbreak, reduced availability of medical services (*e.g*., prenatal and postnatal care), school closures, and feelings of panic spread via social media may adversely affect child health. After the pandemic, socioeconomic challenges such as increased unemployment can also impair child health. Although many underlying channels can explain negative associations between pandemic exposure and child health, the contributions of different pathways to the epidemiological linkage are unknown.

Although the linkages between the pandemic-associated social stress and child development are biologically plausible, few studies have examined these relationships. This paucity may relate to the complexity of measuring dimensions of social stress. Therefore, understanding the effect of social stress on child development requires a comprehensive study design. Novel infectious disease outbreaks are unpredictable, which further exacerbates the challenge of assessments aimed at characterizing the effects of pandemic-associated social stressors. Furthermore, secondary impacts from the pandemic are largely unknown. Some social stressors produce only transient effects in adults and thus are usually overlooked. However, childhood stressors often have long-term irreversible impacts, including the promotion of adult diseases. Thus, to protect child health, long-term views and timely actions are required.

Even though COVID-19 has caused worldwide social stress^9^ and adversely affected maternal and infant health^27,28^, the long-term impacts on child development have not been reported due to the lag between outcomes and exposure. However, according to data from the SARS pandemic, it is plausible that impacts on child growth, and cognitive and physiological development, may occur as a result of exposure to the COVID-19 pandemic. Therefore, the development of public interventions for young children are warranted. For instance, educational efforts to explain the purpose of wearing masks may mitigate stress^16^. School closures should be applied cautiously^6^ and psychological counseling should be provided for children and their caregivers during isolation. Our findings also suggest that preschoolers (3-year-olds) were most sensitive to SARS, and similar hypotheses for COVID-19 should be examined. Furthermore, the differences between SARS and COVID-19 should be noted. As we described in our study, the developmental delay effects were enhanced by pandemic size (Figure 2). Therefore, it is reasonable to assume more severe child-health impacts from COVID-19, as compared to SARS.

We examined cohort effects for children with shared SARS pandemic experiences. Since the long-term trend in child health is controlled in our main models, the estimated effects are attributable to the discontinuous signals at the beginning of 2003, such as the SARS outbreak. Our sensitivity analysis showed that the estimated effect was also spatially correlated with severity of SARS (Figure 2), which further increases the reliability of our findings. However, the models may be challenged by the existence of other events that coincide with SARS. Although such a coincidence is unlikely, it limits the causality of our findings.

There are three additional limitations of our study. First, because of the retrospective design, this study could not avoid survival bias or recall bias, which may have led to underestimation of SARS-related effects on child health. Second, the CFPS cohort was designed for a general purpose, not specifically for child health assessments. Although SARS-associated social stress could impact many dimensions of child development, we could only examine the available measures and may have neglected more important measures. Third, we evaluated the exposure to SARS using simple binary indicators. Since the severity of SARS varied spatially and temporally, our findings might also be biased due to exposure misclassifications. The above limitations could be addressed with a prospective study design. Therefore, a cohort focusing on child development under the COVID-19 pandemic should be urgently planned.

## Conclusion

Our retrospective analysis examined links between SARS pandemic exposure and child health outcomes and found that exposure was associated with delayed developmental milestones and reduced body weight. The associations were strongest in SARS hotspots. Our findings suggest that social stressors associated with novel infectious diseases can impair child development. During the current COVID-19 outbreak, studies and interventions aimed at characterizing and addressing child health effects should be immediately initiated.

## Data Availability

The study is based on public data, which can be accessed from the CFPS website (http://www.isss.pku.edu.cn/cfps/index.htm).

## Author contributions

Y.F., Q.W. and T.X. prepared the data; Y.F., and H.W. analyzed the data; Q.W., X.Z., Y.Z. and B.W. interpreted the results; Y.F. H.W. and T.X. prepared the first draft; T.X. and T.Z. designed the study. All co-authors revised the manuscript together.

## Acknowledgement

We thank the Institute of Social Science Survey, Peking University provided the China Family Panel Studies data; The corresponding author (T.X.) thanks his daughter Xue, Manshu, who was born in Dec. 13^th^, 2019 and had to stay at home for her early developmental stage because of COVID-19. Her experience inspired this study. T.X. also thanks the early career funding from Peking University Health Science Center.

## Supplemental materials

**Table S1.** Associations between SARS and delayed milestones, estimated by different models.

| Milestone | Exposure | Hazard ratio (95% confidence intervals) of developmental delay |  |  |
| --- | --- | --- | --- | --- |
|  |  | Model 1* | Model 2** | Model 3*** |
| Walking independently | SARS<sub>maternal</sub> | 0.946 (0.780, 1.148) | 0.967 (0.798, 1.174) | 1.011 (0.832, 1.229) |
|  | SARS<sub>child</sub> | 3.343 (2.863, 3.903) | 3.323 (2.844, 3.882) | 3.168 (2.710, 3.703) |
| Saying a complete sentence | SARS<sub>maternal</sub> | 0.900 (0.743, 1.091) | 0.906 (0.748, 1.099) | 0.905 (0.746, 1.098) |
|  | SARS<sub>child</sub> | 4.066 (3.571, 4.629) | 4.038 (3.548, 4.597) | 3.984 (3.500, 4.534) |
| Counting from 1 to 10 | SARS<sub>maternal</sub> | 1.021 (0.837, 1.245) | 1.022 (0.838, 1.246) | 1.029 (0.843, 1.256) |
|  | SARS<sub>child</sub> | 5.207 (4.705, 5.763) | 5.018 (4.534, 5.554) | 4.960 (4.481, 5.489) |
| Undressing him/herself for urination | SARS<sub>maternal</sub> | 0.953 (0.782, 1.161) | 0.953 (0.781, 1.161) | 0.967 (0.792, 1.180) |
|  | SARS<sub>child</sub> | 5.685 (5.103, 6.332) | 5.600 (5.028, 6.237) | 5.569 (5.000, 6.203) |

\* Model 1: adjustments of temporal trend and spatial random effect;
\*\* Model 2: model 1 + adjustments of sex, ethnicity, and residence;
\*\*\* Model 3 (fully-adjusted model): model 2 + adjustments of gestational length, birthweight and breastfeeding duration.

**Table S2.**
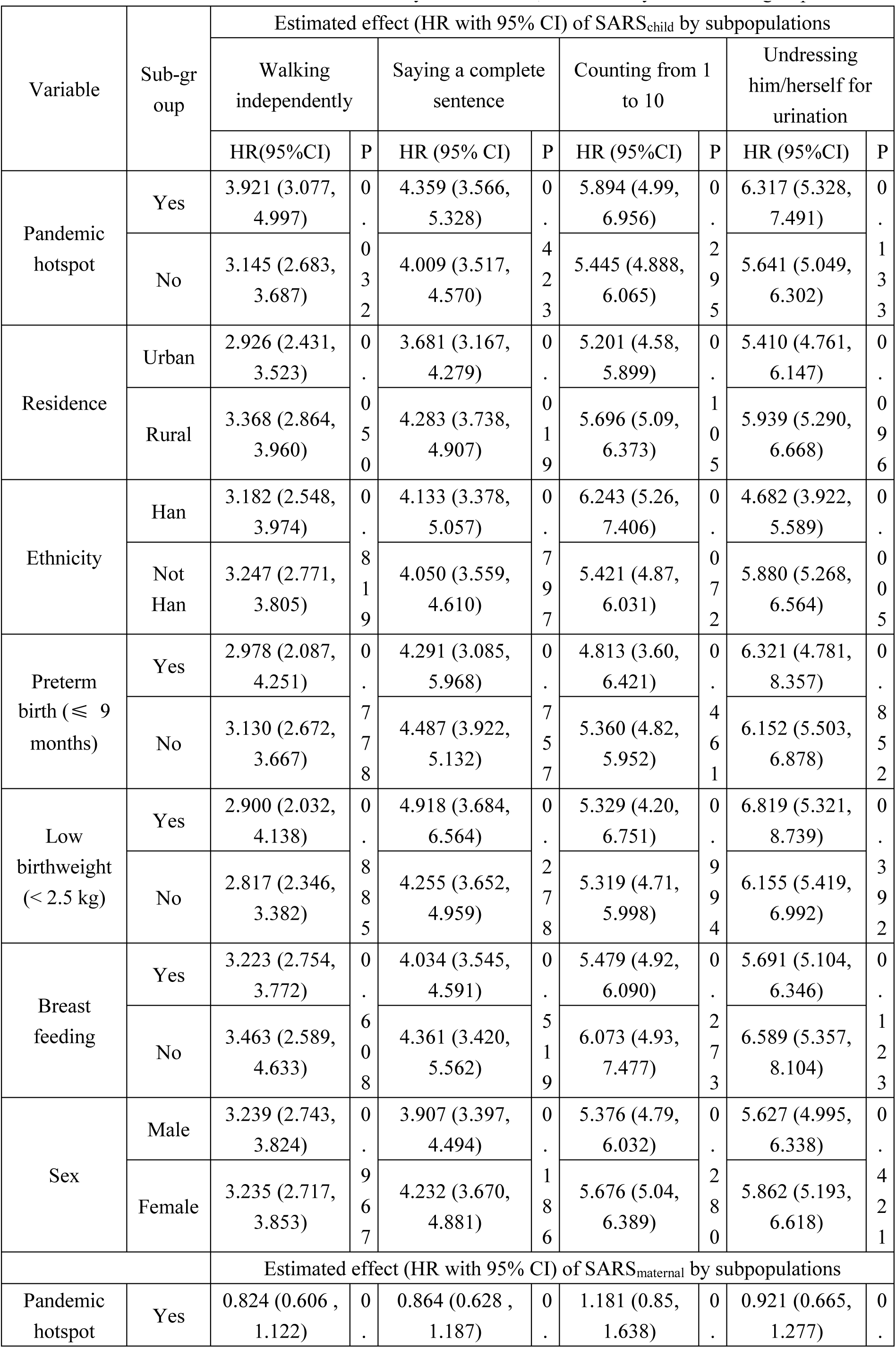
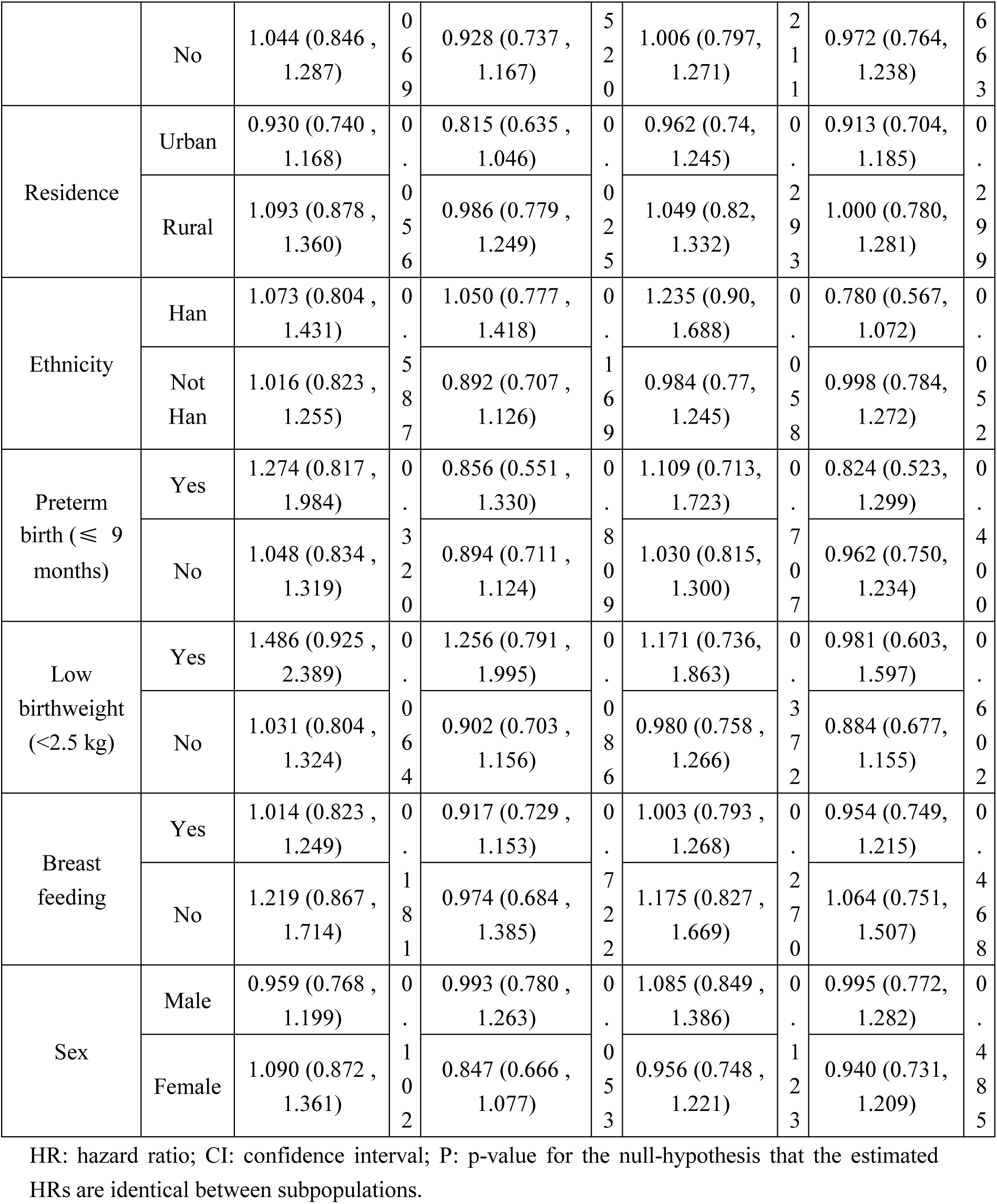
Associations between SARS and delayed milestones, estimated by different subgroups.

**Table S3.** The enhanced association between SARS and delayed milestone by continuous indicators for size of the pandemic.

| Outcome | Enhancement effect* by $x$ (95% CI); P-value | |
| --- | --- | --- |
| | $x$ = number of deaths | $x$ = number of cases |
| Walking independently | 0.46% (-0.02%, 0.94%); P=0.062 | 0.24% (-0.06%, 0.54%); P=0.114 |
| Saying a complete sentence | 0.36% (-0.06%, 0.79%); P=0.096 | 0.26% (-0.01%, 0.53%); P=0.061 |
| Counting from 1 to 10 | 0.31% (-0.05%, 0.68%); P=0.092 | 0.12% (-0.10%, 0.35%); P=0.282 |
| Undressing him/herself for urination | 0.45% (0.08%, 0.82%); P=0.016 | 0.32% (0.08%, 0.55%); P=0.008 |

\* Enhancement effect: the increase in hazard ratio (%) per 10% increment in the number of SARS deaths or cases.

**Table S4.**
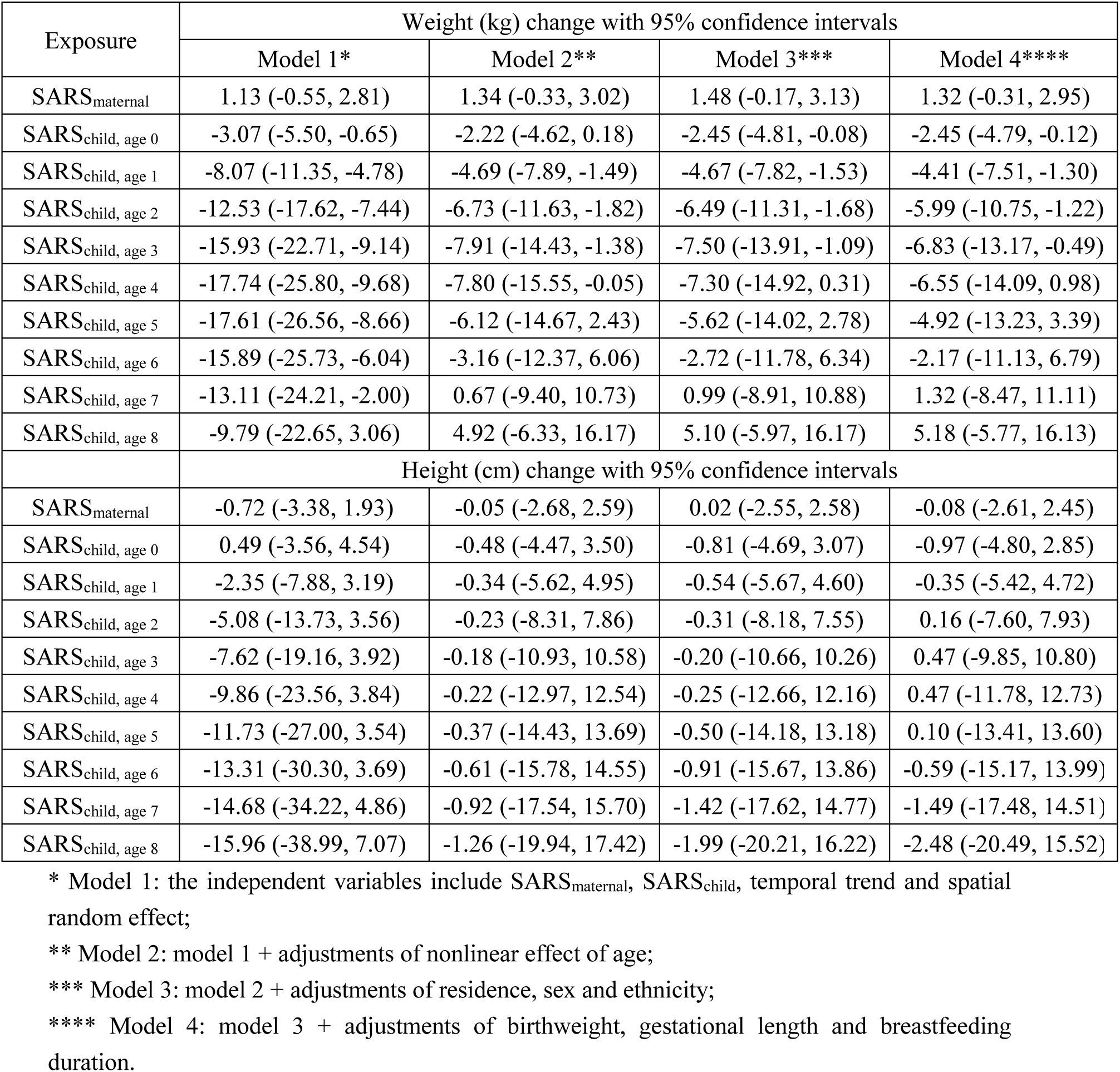
The estimated associations between SARS and body weight/height by different models.

**Table S5.** The estimated associations between SARS and body weight/height by subpopulations.

|  | Weight change (cm) |  | Height change (cm) |  |
| --- | --- | --- | --- | --- |
|  | Pandemic hotspot |  | Pandemic hotspot |  |
|  | No | Yes | No | Yes |
| SARS<sub>maternal</sub> | 1.29 (-0.35, 2.93) | 1.44 (-0.85, 3.73) | -0.22 (-2.77, 2.32) | 1.28 (-2.24, 4.80) |
| SARS<sub>child, age 0</sub> | -2.47 (-4.83, -0.12) | -2.55 (-5.25, 0.15) | -1.08 (-4.90, 2.75) | -0.16 (-4.53, 4.20) |
| SARS<sub>child, age 1</sub> | -4.33 (-7.45, -1.22) | -5.02 (-8.24, -1.79) | -0.36 (-5.43, 4.72) | -0.04 (-5.32, 5.25) |
| SARS<sub>child, age 2</sub> | -5.83 (-10.60, -1.07) | -7.05 (-11.86, -2.25) | 0.25 (-7.52, 8.01) | 0.05 (-7.81, 7.91) |
| SARS<sub>child, age 3</sub> | -6.61 (-12.95, -0.27) | -8.24 (-14.62, -1.86) | 0.62 (-9.71, 10.95) | 0.04 (-10.36, 10.45) |
| SARS<sub>child, age 4</sub> | -6.31 (-13.84, 1.22) | -8.15 (-15.72, -0.57) | 0.64 (-11.62, 12.89) | -0.10 (-12.42, 12.23) |
| SARS<sub>child, age 5</sub> | -4.69 (-12.99, 3.62) | -6.49 (-14.84, 1.85) | 0.23 (-13.28, 13.74) | -0.40 (-13.96, 13.16) |
| SARS<sub>child, age 6</sub> | -1.98 (-10.94, 6.97) | -3.56 (-12.54, 5.43) | -0.52 (-15.11, 14.06) | -0.83 (-15.46, 13.80) |
| SARS<sub>child, age 7</sub> | 1.44 (-8.35, 11.23) | 0.23 (-9.60, 10.06) | -1.51 (-17.51, 14.49) | -1.36 (-17.44, 14.72) |
| SARS<sub>child, age 8</sub> | 5.22 (-5.73, 16.18) | 4.44 (-6.59, 15.47) | -2.62 (-20.63, 15.39) | -1.93 (-20.09, 16.23) |
|  | Residence |  | Residence |  |
|  | Rural | Urban | Rural | Urban |
| SARS<sub>maternal</sub> | 0.60 (-1.09, 2.29) | 1.98 (0.31, 3.65) | -0.65 (-3.33, 2.03) | 0.39 (-2.23, 3.01) |
| SARS<sub>child, age 0</sub> | -3.11 (-5.68, -0.55) | -2.65 (-4.99, -0.30) | -1.18 (-5.06, 2.69) | -1.63 (-5.96, 2.69) |
| SARS<sub>child, age 1</sub> | -5.81 (-8.99, -2.63) | -4.77 (-7.87, -1.67) | -0.67 (-5.76, 4.42) | -1.33 (-6.59, 3.93) |
| SARS<sub>child, age 2</sub> | -8.12 (-12.89, -3.34) | -6.53 (-11.26, -1.79) | -0.24 (-8.01, 7.53) | -1.07 (-8.93, 6.79) |
| SARS<sub>child, age 3</sub> | -9.65 (-15.98, -3.31) | -7.56 (-13.85, -1.26) | 0.02 (-10.32, 10.35) | -0.90 (-11.32, 9.51) |
| SARS<sub>child, age 4</sub> | -10.01 (-17.53, -2.50) | -7.49 (-14.97, -0.01) | 0.02 (-12.24, 12.28) | -0.88 (-13.21, 11.46) |
| SARS<sub>child, age 5</sub> | -8.96 (-17.24, -0.67) | -6.09 (-14.34, 2.17) | -0.29 (-13.80, 13.22) | -1.02 (-14.59, 12.55) |
| SARS<sub>child, age 6</sub> | -6.74 (-15.67, 2.20) | -3.59 (-12.50, 5.31) | -0.86 (-15.44, 13.73) | -1.30 (-15.95, 13.35) |
| SARS<sub>child, age 7</sub> | -3.74 (-13.54, 6.05) | -0.37 (-10.10, 9.37) | -1.59 (-17.60, 14.41) | -1.68 (-17.80, 14.44) |
| SARS<sub>child, age 8</sub> | -0.36 (-11.37, 10.65) | 3.22 (-7.67, 14.12) | -2.42 (-20.45, 15.61) | -2.10 (-20.34, 16.14) |
|  | Ethnicity |  | Ethnicity |  |
|  | Han | Not Han | Han | Not Han |
| SARS<sub>maternal</sub> | 1.44 (-0.21, 3.08) | 1.14 (-1.04, 3.32) | -0.04 (-2.61, 2.52) | 0.78 (-2.70, 4.27) |
| SARS<sub>child, age 0</sub> | -1.32 (-4.11, 1.47) | -2.34 (-4.68, -0.00) | 2.55 (-2.14, 7.25) | -0.60 (-4.43, 3.24) |
| SARS<sub>child, age 1</sub> | -2.03 (-5.30, 1.24) | -4.14 (-7.24, -1.04) | 2.72 (-2.69, 8.12) | -0.08 (-5.15, 4.99) |
| SARS<sub>child, age 2</sub> | -2.50 (-7.34, 2.34) | -5.57 (-10.32, -0.82) | 2.89 (-5.03, 10.82) | 0.35 (-7.41, 8.11) |
| SARS<sub>child, age 3</sub> | -2.47 (-8.90, 3.95) | -6.28 (-12.60, 0.04) | 3.10 (-7.38, 13.57) | 0.61 (-9.70, 10.92) |
| SARS<sub>child, age 4</sub> | -1.71 (-9.32, 5.90) | -5.90 (-13.41, 1.61) | 3.34 (-9.04, 15.73) | 0.62 (-11.62, 12.85) |
| SARS<sub>child, age 5</sub> | -0.04 (-8.41, 8.32) | -4.18 (-12.47, 4.10) | 3.64 (-9.96, 17.24) | 0.31 (-13.17, 13.80) |
| SARS<sub>child, age 6</sub> | 2.36 (-6.64, 11.37) | -1.37 (-10.31, 7.56) | 3.98 (-10.69, 18.65) | -0.25 (-14.81, 14.32) |
| SARS<sub>child, age 7</sub> | 5.26 (-4.60, 15.13) | 2.16 (-7.60, 11.92) | 4.35 (-11.81, 20.51) | -0.97 (-16.95, 15.00) |
| SARS<sub>child, age 8</sub> | 8.41 (-2.71, 19.52) | 6.06 (-4.86, 16.98) | 4.73 (-13.61, 23.08) | -1.79 (-19.77, 16.20) |
|  | Sex |  | Sex |  |
|  | Female | Male | Female | Male |
| SARS<sub>maternal</sub> | 1.19 (-0.51, 2.89) | 1.56 (-0.17, 3.29) | 0.77 (-1.91, 3.45) | -0.88 (-3.55, 1.79) |
| SARS<sub>child, age 0</sub> | -4.51 (-7.23, -1.78) | -3.16 (-5.53, -0.78) | -1.98 (-6.46, 2.50) | -1.15 (-5.05, 2.74) |
| SARS<sub>child, age 1</sub> | -6.40 (-9.62, -3.18) | -5.03 (-8.14, -1.92) | -0.96 (-6.23, 4.32) | -0.38 (-5.46, 4.70) |
| SARS<sub>child, age 2</sub> | -7.97 (-12.78, -3.16) | -6.56 (-11.32, -1.80) | -0.23 (-8.09, 7.63) | 0.23 (-7.53, 8.00) |
| SARS<sub>child, age 3</sub> | -8.91 (-15.32, -2.51) | -7.40 (-13.74, -1.06) | -0.08 (-10.52, 10.35) | 0.51 (-9.82, 10.83) |
| SARS<sub>child, age 4</sub> | -8.91 (-16.51, -1.31) | -7.19 (-14.73, 0.34) | -0.81 (-13.18, 11.55) | 0.27 (-11.98, 12.53) |
| SARS<sub>child, age 5</sub> | -7.75 (-16.12, 0.61) | -5.72 (-14.03, 2.58) | -2.61 (-16.20, 10.98) | -0.58 (-14.08, 12.92) |
| SARS<sub>child, age 6</sub> | -5.65 (-14.65, 3.34) | -3.21 (-12.16, 5.74) | -5.29 (-19.93, 9.35) | -1.94 (-16.51, 12.63) |
| SARS<sub>child, age 7</sub> | -2.92 (-12.75, 6.91) | -0.01 (-9.78, 9.77) | -8.55 (-24.64, 7.54) | -3.64 (-19.62, 12.34) |
| SARS<sub>child, age 8</sub> | 0.13 (-10.91, 11.17) | 3.54 (-7.39, 14.48) | -12.10 (-30.29, 6.09) | -5.50 (-23.49, 12.49) |
|  | Preterm birth |  | Preterm birth |  |
|  | No | Yes | No | Yes |
| SARS<sub>maternal</sub> | 1.69 (-0.05, 3.43) | 1.16 (-0.56, 2.87) | -0.01 (-2.72, 2.71) | 0.04 (-2.64, 2.71) |
| SARS<sub>child, age 0</sub> | -2.22 (-5.02, 0.57) | -2.41 (-4.82, -0.01) | 0.56 (-4.02, 5.14) | -0.41 (-4.35, 3.53) |
| SARS<sub>child, age 1</sub> | -3.81 (-7.09, -0.54) | -4.23 (-7.37, -1.10) | 0.01 (-5.36, 5.38) | -0.18 (-5.30, 4.93) |
| SARS<sub>child, age 2</sub> | -5.05 (-9.89, -0.22) | -5.69 (-10.46, -0.93) | -0.47 (-8.37, 7.43) | -0.00 (-7.79, 7.78) |
| SARS<sub>child, age 3</sub> | -5.59 (-12.00, 0.81) | -6.44 (-12.78, -0.09) | -0.81 (-11.24, 9.63) | 0.09 (-10.25, 10.43) |
| SARS<sub>child, age 4</sub> | -5.09 (-12.68, 2.50) | -6.10 (-13.64, 1.43) | -0.93 (-13.28, 11.42) | 0.05 (-12.21, 12.31) |
| SARS<sub>child, age 5</sub> | -3.30 (-11.65, 5.05) | -4.46 (-12.77, 3.85) | -0.80 (-14.38, 12.77) | -0.15 (-13.66, 13.36) |
| SARS<sub>child, age 6</sub> | -0.47 (-9.46, 8.52) | -1.73 (-10.69, 7.23) | -0.46 (-15.10, 14.18) | -0.48 (-15.06, 14.11) |
| SARS<sub>child, age 7</sub> | 3.05 (-6.79, 12.90) | 1.71 (-8.07, 11.50) | 0.02 (-16.10, 16.14) | -0.90 (-16.91, 15.11) |
| SARS<sub>child, age 8</sub> | 6.93 (-4.14, 18.00) | 5.52 (-5.44, 16.47) | 0.57 (-17.69, 18.83) | -1.36 (-19.39, 16.68) |
|  | Low birthweight |  | Low birthweight |  |
|  | No | Yes | No | Yes |
| SARS<sub>maternal</sub> | 1.37 (-0.26, 3.00) | -0.80 (-3.39, 1.78) | -0.13 (-2.66, 2.40) | 3.26 (-1.84, 8.37) |
| SARS<sub>child, age 0</sub> | -3.62 (-6.54, -0.70) | -2.53 (-4.86, -0.19) | -2.54 (-7.34, 2.25) | -0.91 (-4.74, 2.91) |
| SARS<sub>child, age 1</sub> | -4.88 (-8.20, -1.56) | -4.43 (-7.54, -1.32) | -1.87 (-7.25, 3.51) | -0.24 (-5.31, 4.84) |
| SARS<sub>child, age 2</sub> | -5.82 (-10.67, -0.96) | -5.97 (-10.74, -1.21) | -1.26 (-9.17, 6.64) | 0.33 (-7.44, 8.10) |
| SARS<sub>child, age 3</sub> | -6.12 (-12.54, 0.31) | -6.78 (-13.12, -0.44) | -0.79 (-11.27, 9.70) | 0.68 (-9.66, 11.02) |
| SARS<sub>child, age 4</sub> | -5.46 (-13.08, 2.15) | -6.49 (-14.02, 1.05) | -0.50 (-12.93, 11.92) | 0.71 (-11.56, 12.97) |
| SARS<sub>child, age 5</sub> | -3.64 (-12.02, 4.73) | -4.85 (-13.16, 3.46) | -0.46 (-14.10, 13.19) | 0.34 (-13.18, 13.85) |
| SARS<sub>child, age 6</sub> | -0.87 (-9.88, 8.15) | -2.11 (-11.07, 6.85) | -0.60 (-15.31, 14.10) | -0.36 (-14.95, 14.23) |
| SARS<sub>child, age 7</sub> | 2.55 (-7.32, 12.42) | 1.36 (-8.43, 11.15) | -0.88 (-17.06, 15.31) | -1.28 (-17.28, 14.73) |
| SARS<sub>child, age 8</sub> | 6.28 (-4.84, 17.41) | 5.20 (-5.76, 16.15) | -1.22 (-19.59, 17.15) | -2.30 (-20.31, 15.71) |
|  | Breastfeeding |  | Breastfeeding |  |
|  | No | Yes | No | Yes |
| SARS<sub>maternal</sub> | 2.10 (-0.23, 4.44) | 1.28 (-0.36, 2.92) | -1.30 (-5.09, 2.49) | 0.03 (-2.51, 2.56) |
| SARS<sub>child, age 0</sub> | -1.07 (-5.09, 2.95) | -1.82 (-4.62, 0.97) | 0.41 (-6.14, 6.96) | -0.31 (-4.88, 4.26) |
| SARS<sub>child, age 1</sub> | -2.73 (-6.56, 1.09) | -3.58 (-6.87, -0.28) | 1.59 (-4.68, 7.87) | 0.65 (-4.73, 6.04) |
| SARS<sub>child, age 2</sub> | -4.18 (-9.44, 1.09) | -5.04 (-9.94, -0.14) | 2.38 (-6.23, 10.99) | 1.36 (-6.64, 9.36) |
| SARS<sub>child, age 3</sub> | -5.17 (-12.08, 1.75) | -5.92 (-12.41, 0.58) | 2.37 (-8.88, 13.63) | 1.55 (-9.03, 12.13) |
| SARS<sub>child, age 4</sub> | -5.48 (-13.56, 2.61) | -5.92 (-13.60, 1.76) | 1.18 (-11.95, 14.30) | 0.95 (-11.53, 13.44) |
| SARS<sub>child, age 5</sub> | -4.96 (-13.67, 3.76) | -4.84 (-13.26, 3.58) | -1.47 (-15.62, 12.68) | -0.60 (-14.28, 13.08) |
| SARS<sub>child, age 6</sub> | -3.76 (-13.10, 5.58) | -2.89 (-11.96, 6.17) | -5.31 (-20.55, 9.93) | -2.93 (-17.69, 11.84) |
| SARS<sub>child, age 7</sub> | -2.11 (-12.63, 8.40) | -0.36 (-10.35, 9.63) | -9.94 (-27.27, 7.40) | -5.78 (-22.15, 10.59) |
| SARS<sub>child, age 8</sub> | -0.24 (-12.66, 12.19) | 2.47 (-8.89, 13.83) | -14.96 (-35.63, 5.71) | -8.90 (-27.65, 9.85) |

